# Optimal age targeting for pneumococcal vaccination in older adults; a modelling study

**DOI:** 10.1101/2022.11.07.22282045

**Authors:** Deus Thindwa, Samuel Clifford, Jackie Kleynhans, Anne von Gottberg, Sibongile Walaza, Susan Meiring, Todd D Swarthout, Elizabeth Miller, Peter McIntyre, Nick Andrews, Zahin Amin-Chowdhury, Norman Fry, Kondwani Jambo, Neil French, Samanta Cristine Grassi Almeida, Shamez Ladhani, Robert S Heyderman, Cheryl Cohen, Maria Cristina de Cunto Brandileone, Stefan Flasche

**Affiliations:** Centre for Mathematical Modelling of Infectious Diseases, London School of Hygiene & Tropical Medicine, London, UK; Department of Infectious Disease Epidemiology London School of Hygiene & Tropical Medicine, London, UK; Malawi Liverpool Wellcome Research Programme, Blantyre, Malawi; Centre for Respiratory Diseases and Meningitis, National Institute for Communicable Diseases of the National Health Laboratory Service, Johannesburg, South Africa; School of Public Health, University of the Witwatersrand, Johannesburg, South Africa; School of Pathology, University of the Witwatersrand, Johannesburg, South Africa; Division of Infection and Immunity, University College London, London, UK; Immunisation and Countermeasures Division, UK Health Security Agency, London, UK; University of Otago, Dunedin, New Zealand; Department of Clinical Sciences, Liverpool School of Tropical Medicine, Liverpool, UK; Institute of Infection, Veterinary and Ecological Sciences, University of Liverpool, UK; National Laboratory for Meningitis and Pneumococcal Infections, Laboratory for Meningitis, Pneumonia and Pneumococcal Infection, Centre of Bacteriology, Brazil

## Abstract

**Background:** Invasive pneumococcal disease (IPD) risk increases with age for older adults whereas the population size benefiting from pneumococcal vaccines and the robustness of the immunogenic response to vaccination decline. This poses a conundrum for identifying the optimal age for vaccination. We estimate how demographics, vaccine efficacy/effectiveness (VE), and waning VE impact on optimal age for single-dose pneumococcal vaccination.

**Methods:** Age- and vaccine-serotype-stratified IPD incidence from routine surveillance of adults ≥55 years old (y) in Brazil, England, Blantyre (Malawi), and South Africa, ≥4-years after infant-pneumococcal vaccine introduction and before 2020, was used to parameterise exponential growth models of increasing IPD risk with age. A piecewise-constant model estimated VE and waning VE from prior studies. All estimates were then combined in a cohort model to assess the vaccine preventable IPD burden of delivering 13-, 15-, 20-valent conjugate or 23-valent polysaccharide vaccines at various ages.

**Findings:** In Brazil, Malawi, South Africa and England 51%, 51%, 54% and 39% of adults older than 55y were younger than 65y old. A smaller share of IPD was reported among adults <65y old in England (4,657 annual cases; 20% <65y) compared to Brazil (186; 45%), Malawi (4; 63%), or South Africa (134, 48%). PCV13- and PPV23-serotypes were estimated to cause 39% and 85% of IPD cases on average across settings. Vaccination at 55y in Brazil, Malawi, and South Africa, and at 70y in England had the greatest potential for IPD prevention with 38%, 31%, 27%, 8% of IPD preventable if using PCV13, 44%, 32%, 30%, 13% with PCV15, 65%, 69%, 53%, 37% with PCV20, and 30%, 29%, 20%, 14% with PPV23, respectively. Vaccination efficiency or cost effective use was optimal in 60-65y in Brazil, Malawi and South Africa, and in 80-85y in England.

**Interpretation:** In low/middle-income countries, pneumococcal vaccines may prevent a substantial proportion of the residual IPD burden if administered earlier in adulthood than is typical in high-income countries.

**Funding:** UK National Institute for Health and Care Research.

**Research in context:** *Evidence before this study:* A recent review conducted for the World Health Organisation (WHO) Scientific Advisory Group of Experts (SAGE) working group on pneumococcal vaccines reviewed the evidence on the efficacy, effectiveness and potential impact of pneumococcal vaccines in older adults. At the time, little evidence was identified to support recommendations for the age of pneumococcal vaccination in older adults, particularly in LMICs. We did an updated search of PubMed, Medline, Embase and Web of Science for studies on age targeting for pneumococcal vaccination in older adults published from Jan 1, 2003 to June 30, 2022, using the search terms “(optimal age targeting OR age targeting OR timeliness) AND (pneumococcal vaccination OR pneumococcal vaccines) AND (adults OR older adults OR elderly)”. We identified one study on the role of timeliness in the cost-effectiveness of older adult vaccination in Australia. This study explored the impact of the timeliness of pneumococcal conjugate vaccination (PCV) in older adults and found that more hospitalisations and deaths can be prevented if PCV13 was given to 70y old than as recommended at 65y. On the other hand, most low-income countries (LICs) and middle-income countries (MICs) do not have routine older adult pneumococcal vaccination programmes, despite often less indirect protection from childhood PCV programmes and high burden of vaccine preventable disease. The implications of differences in population demographics and risk for IPD compared to high income countries on the optimal age for pneumococcal vaccination in LICs/MICs are unknown.

*Added value of this study:* This multicountry study uses data from long-standing older adult pneumococcal surveillance programmes at the national/sub-national level, to explore the optimal age-targeting for a single-dose pneumococcal vaccination against vaccine-serotype-IPD in older adults living without human immunodeficiency virus in LIC (Malawi), MICs (Brazil, South Africa), and HIC (England). We show that despite mature pneumococcal infant vaccination programmes there is a substantial burden of vaccine preventable pneumococcal disease remaining in the four countries considered. Vaccinating at 55y in the LICs/MICs was optimal whereas vaccinating at 70y prevented most IPD cases in England than vaccinating at other ages. While the number of older adults needed to vaccinate to prevent an IPD case may be higher for programmes vaccinating older individuals, vaccination efficiency or vaccine cost effective use was optimal in 60-65y in LICs/MICs, and in 80-85y in England.

*Implications of all available evidence:* LICs/MICs with a mature infant immunisation programme for pneumococci and a remaining high burden of vaccine preventable pneumococcal disease in older adults may consider implementation of a vaccination programme for older adults, albeit at a younger age than typical in HICs. Affordability and cost-effectiveness of an adult vaccination programme as well as the indirect effects following potential changes to higher valency infant PCV vaccination programmes will also need to be considered.

## Introduction

*Streptococcus pneumoniae* (pneumococcus) is a major global cause of childhood mortality [1,2], but also causes a high burden of disease among older adults [2,3]. Two vaccines have been used to prevent pneumococcal disease in older adults ≥55 years-old (y): a 13-valent pneumococcal conjugate vaccine (PCV13) and a 23-valent pneumococcal polysaccharide vaccine (PPV23) [4]. Recently, 15- and 20-valent PCVs (PCV15, PCV20) have also been licensed and recommended for older adults in the United States [5,6].

Although routine infant PCV programmes have generated indirect protection against vaccine-serotype (VT) pneumococcal disease among older adults [7,8], a substantial disease burden remains, composed of serotypes not targeted by childhood PCV programmes and residual circulation of VT [2,9]. Among high-income countries (HICs) with a mature infant-PCV13 programme, PCV13- and PPV23-targeted serotypes caused about 15% and 42%, respectively, of invasive pneumococcal disease (IPD) in older adults [2,10]. Routine infant-PCV programmes in many low- and middle-income countries (LICs and MICs) have often led to less pronounced herd effects with continued circulation of VTs especially in the unvaccinated adult population [11,12], who happen to have no access to routine pneumococcal vaccination [13].

The recommended age for pneumococcal vaccination in older adults in HICs is typically either at 60y or 65y [4,14]. However, yet only a single study has assessed the age at which the most gain from such programme is seen (in the Australian context) [15]. As the risk of severe pneumococcal disease increases with age [10,15], waning of protection from vaccination early in older adulthood risks disease at ages of highest disease incidence, whereas vaccination late in older adulthood cannot address the substantial disease burden among a large pool of susceptible older adults who have not yet been vaccinated.

In this modelling study, we explore the optimal age-targeting for a single-dose pneumococcal vaccination against VT-IPD in older adults living without human immunodeficiency virus (HIV) in Brazil, England, Malawi, and South Africa.

## Methods

### Study sites

We included England (HIC), Brazil and South Africa (MICs), and Blantyre in Malawi (LIC) in this analysis, to explore the optimal age-targeting for pneumococcal vaccination, as all have long-standing adult pneumococcal surveillance programmes at the national or sub-national level [9,16–18]. More details on the respective IPD surveillance systems are in the appendix (Text S1). Infant-PCV13 has been in use since 2011 in South Africa (two primary doses and booster (2+1) schedule) and Malawi (3+0 schedule), with estimated 90-95% vaccination coverage [11,18]. In England, routine PCV13 immunisation programme with 92% booster dose coverage has been in place since 2010, initially using a 2+1 schedule, switching to 1+1 schedule in 2020 [19]. In Brazil, a routine infant-PCV10 programme was implemented in 2010 with a 3+1 schedule, switched to a 2+1 in 2016, with average coverage of 94% between 2015 and 2017 [20].

In England, PPV23 has been recommended for risk groups since 1992, and for all adults aged ≥65y since 2003, with single-dose coverage rising to 70% in 2018 [17,21]. In Brazil, PPV23 is not included in the national immunisation programme but has been recommended for use since the 1980s, and is available free of charge at the centers of special immunobiological for people above 2 years old including institutionalised older adult, and thus coverage of <1% among all older adults above 60y [22]. In South Africa, although adult pneumococcal vaccination is recommended, there is no routine programme nor nationally accepted guidelines [23]. In Malawi, neither routine adult pneumococcal vaccination programme nor national guidelines exist [13].

### Population demographics

The population demographics for adults ≥55y were obtained in annual age strata from population censuses conducted in 2010 for Brazil [24], 2011 England (updated in 2017) [25], 2018 Malawi [26], and 2011 South Africa [27]. To align with IPD surveillance timelines in respective countries, annual age population censuses were projected at a constant growth rate of 0·8% during 2010-2016 in Brazil and 1·3% during 2011-2016 in South Africa [28,29]. For England and Malawi, 2017 and 2018 populations already aligned with IPD surveillance time. Annual age population estimates were smoothed to reduce demographic stochasticity in downstream results by applying a 5-year moving average [30].

### Invasive pneumococcal disease burden

For each site we used data on IPD burden in older adults in the presence of a mature infant-PCV programme, defined as at least 4 years after infant-PCV introduction, to avoid inclusion of ongoing changes in IPD burden attributable to indirect effects from the childhood programme [31]. The number of IPD cases caused by all serotypes, and serotypes targeted by PCV13, PCV15, PCV20 and PPV23 in annual age strata (55y to 85y+) were obtained from laboratory-based surveillance in each country (Fig S1). We calculated age and serotype distribution and proportionally inflated estimates by the number of IPD cases without serotyping available to correct for proportion serotyped. In England, where PPV23 has been in use in 65y+ at 70% uptake, we back-inflated the number of IPD cases in order to correct for the fact that in the absence of PPV23 vaccination, IPD incidence would be higher, especially in ages eligible for PPV23 vaccination. Details about back-inflation calculation are in the appendix (Text S2). Age aggregated (55-59y, 60-64y, 65-69y, 70-74y, 75-79y, 80-84y and 85y+) and serotype-specific IPD incidence was calculated by dividing by the age-group specific population estimates. Due to the small number of reported IPD cases and incomplete serotyping information in Malawi, we only calculated the serotype distribution for all cases and assumed that it was the same in each age group.

The focus of this study was vaccination strategies for older adults living without HIV because adults living with HIV often have independent pneumococcal vaccine recommendations [12]. Thus, for adult HIV prevalence of >10% in South Africa [32], we adjusted IPD incidence estimates for HIV status. For 201 (18·7%) reported IPD cases, HIV status was reported. We took a 30% random subset of IPD cases with known HIV status and estimated a 49.3% proportion of IPD cases without HIV for this subset. We assumed that a similar proportion of IPD cases with unknown HIV status would be without HIV. We further accounted for similar serotype distribution in those with and without HIV. In Malawi, IPD cases could not be stratified by HIV infection status, and thus we used the age-dependent HIV infection rates in the general adult population and applied the relative propensity for IPD in older adults with and without HIV from South Africa to infer the incidence of IPD among older adults without HIV by age.

We modelled the reported age-specific IPD incidence among older adults without HIV per country as a function of age using an exponential growth model. Details about model fitting are in appendix (Text S3). The expected incidence at age *a* is:

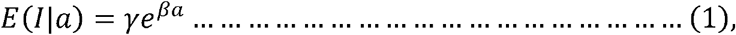

where *E*(*l*|*a*) is the expected IPD incidence at age *a, β* is the growth rate, and *γ* is a constant of proportionality. Uncertainty was obtained by bootstrap sampling 1000 times using the fitted parameter means and covariance matrix and summarisation of uncertainty via 95% quantiles of samples.

### PCV and PPV vaccine efficacy/effectiveness

A previous systematic review identified studies that estimate vaccine efficacy/effectiveness (VE) against IPD from time since vaccination [2]. Four observational studies in HICs estimated VE at two or more time points for PPV23 [17,21,33–35]. One randomised controlled trial estimated VE against IPD and community-acquired pneumonia (CAP) following PCV13 use in older adults and found consistent 75% and 46% respective efficacy for 5 years following vaccination [36,37]. Thus, we assumed that PCVs’ efficacy against IPD would continue to stay stable for five years, and thereafter decline in the same way as the scenario for PPV23. We represented the VE as a function of time since vaccination using piecewise constant models. The piecewise constant functions are shown in appendix (Fig S2, Fig S3). The VE was sampled using bootstrap sampling from a normal distribution centred on the mean VE at that time and standard deviation derived from the reported 95% interval.

Little evidence is available on changes in PPV23 or PCV13 VE by age at administration [15,38]. A UK study found no significant difference in VE of PPV23 when administered to 65y, 75y or 85y adults, although point estimates suggested a decline with age [17]. There is also some evidence that immune response to immunisation is partially impaired later in life [2]. Thus, as a base case we assumed that VE for all formulations was independent of the age of administration, but included a sensitivity analysis assuming that VE was 33% less of initial VE if given to adults aged 65-74y and 44% less of initial VE if given to adults aged 75+y using point estimates of VE of PPV23 against IPD in the 5 years following administration in adults <65y of 54%, decreasing to 36% for 65-74y and 30% for 75y+ [17].

### PCV and PPV vaccine impact model

We developed a cohort model to simulate the risk for IPD in adults >55y in all four countries. Time steps in the model were set at 1 year. Age-dependent all-cause mortality was set to match observed population demographics of older adults living without HIV in all countries, and age-dependent IPD risks were set to match fitted IPD incidence in respective countries and years. We calculated the vaccine preventable number of IPD cases as the lifetime number of cases averted through vaccinating 100% of adults at the specified age for vaccination under the scenarios of age-dependency on initial VE and waning VE. We estimated the efficiency of the alternative strategies by reporting the number of age cohort individuals needed to vaccinate (or number of doses needed to administer to age cohort) in order to prevent a case.

Sensitivity analysis assessed the influence of age-dependent initial VE on vaccine impact. All analyses were conducted using R language v4·1·1, with data and code available through GitHub https://github.com/deusthindwa/optimal.age.targeting.pneumo.vaccines.

### Ethical approval

In Brazil, no patient consent is required since data are obtained through the National Epidemiological Surveillance approved by the Scientific Committee of the Instituto Adolfo Luiz (CTC 61-M/2020). In England, the UK Health Security Agency (UKHSA) has legal permission, provided by Regulation 3 of The Health Service Regulations 2002, to monitor the safety and effectiveness of vaccines for national surveillance of communicable diseases (http://www.legislation.gov.uk/uksi/2002/1438/regulation/3/made). In Malawi, the study protocol was approved by Malawi’s National Health Sciences Research Committee (protocol 867), Kamuzu University of Health Sciences College of Medicine Research Ethics Committee (COMREC) (P·01/08/609 and P·09/09/826), and the University of Liverpool Research Ethics Committee (RETH490). Individual patient informed consent was not required for the use of publicly available anonymised routine samples as per COMREC guidelines 5·6. In South Africa, ethical approval to conduct laboratory-based and enhanced surveillance was obtained from the Health Research Ethics Committee (Human), University of Witwatersrand (M140159) and individual patient consent was obtained for clinical data collection at enhanced surveillance sites. Ethical approval for this study was granted by the London School of Hygiene and Tropical Medicine (25787).

## Results

### Population demographics and IPD burden

Of the 27·7 million (m) older adults (≥55y) in Brazil, 16.5m in England, 66,589 in Blantyre Malawi, and 6·9m in South Africa, the proportion of older adults aged 55y to <65y was 51·3%, 51·0% and 53·8% in Brazil, Malawi and South Africa, substantially higher than the 39·1% in England. During the study period, Brazil (2015-2017), England (2016-2019), Malawi (2016-2019) and South Africa (2015-2018) reported 559, 13,971, 19, 537 IPD cases in adults ≥55y equivalent to an average annual number of cases of 186·3, 4,658·0, 4·8 and 134·3, respectively. Of these in the ≥55y, 44·5%, 19·9%, 62·5%, and 47·9% were in <65y olds. Cases caused by PCV13 serotypes accounted for 61·4% (343/559), 21·0% (2,936/13,971), 41·0% (8/19), and 32·8% (176/537) of all IPD in Brazil, England, Malawi and South Africa, and PPV23 serotypes for 97·9% (547/559), 72·5% (10,126/13,971), 94·8% (18/19), and 73·2% (393/537), respectively (Fig 1).

**Figure 1.**
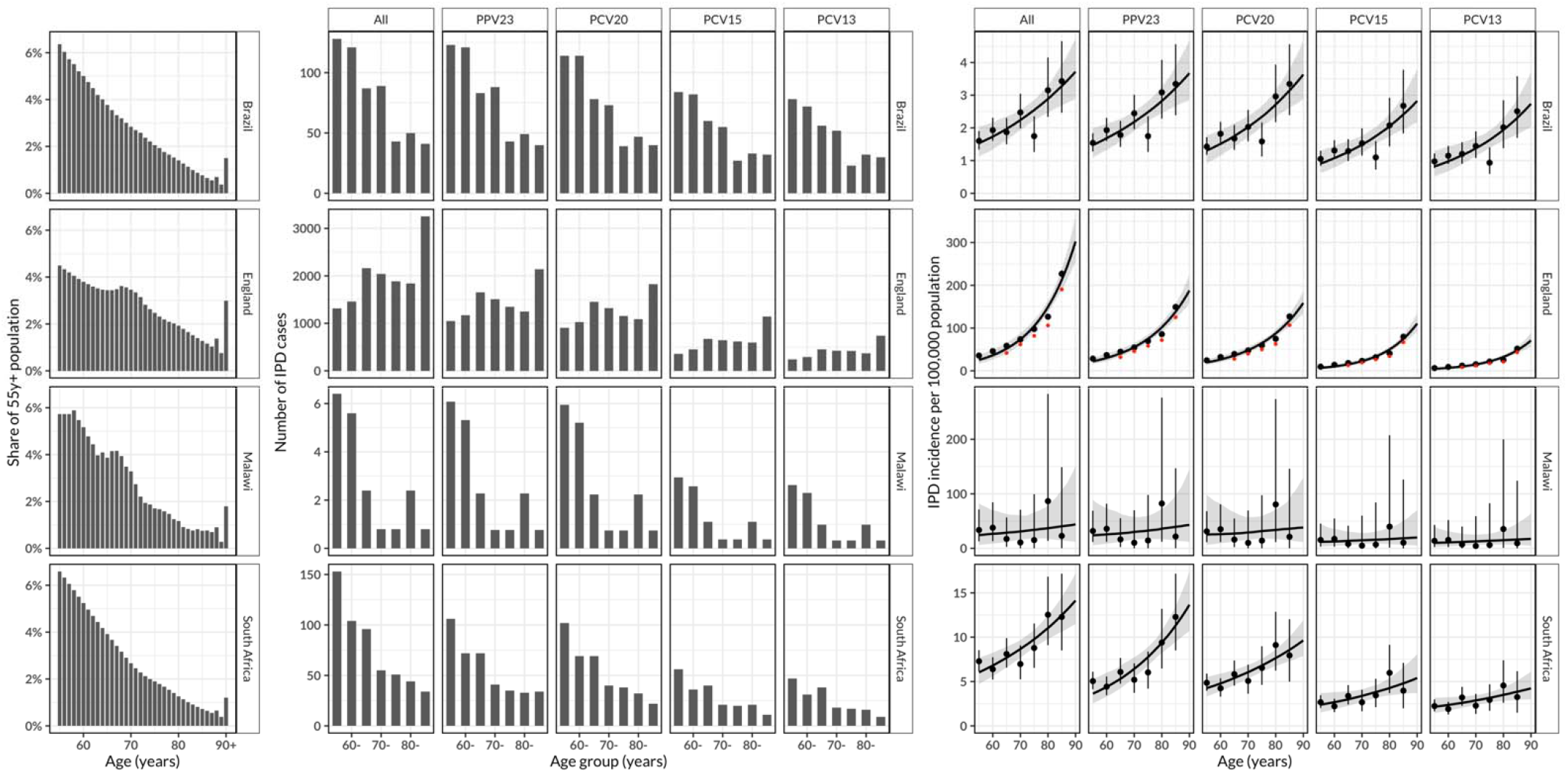
Population demographics and invasive pneumococcal disease (IPD) burden. (Left panel) Among individuals who are aged ≥55y, the proportion/share in annual age groups in Brazil, England, Malawi and South Africa as estimated from their national censuses, based on five years rolling average smoothed population counts to control for demographic stochasticity; (Middle panel) Number of IPD cases in five year age bands in older adults stratified by serotype in Brazil (2015-2017), England (2016-2019), Blantyre Malawi (2016-2019) and South Africa (2015-2018), reported from at least four years post-infant PCV introduction in each country. (Right panel) Serotype-specific reported and predicted IPD incidence per 100,000 population between 55y and 90y in Brazil, England, Malawi and South Africa. The black circle represents estimated IPD cases per 100,000 population, the vertical line through the circle represents a 95% uncertainty interval in estimated IPD case number, the curve line is the exponential model fit and the ribbon represents a bootstrapped 95% confidence interval for the fitted line. The red and black points for England represent estimated IPD cases in the presence and absence of PPV23 vaccination, respectively.

The exponential model fitted the increase in IPD incidence with age well. The estimated IPD incidence in 85y was higher than in 55y olds by 2·48-fold (95% confidence interval [95%CI]: 2·13-2·83) in Brazil, 2·19-fold (95% CI: 0·14-4·51) in Malawi, and 2·25-fold (95% CI: 1·88-2·62) in South Africa. In England the incidence increased more steeply to 11·00-fold (95% CI: 10·90-11·40) higher in 85y than 55y (Fig 1).

While the estimated number of IPD cases declined with age in Brazil, Malawi and South Africa, it increased in England. On the other hand, age-specific IPD incidence of total IPD incidence (age-scaled IPD incidence) increased with age in all settings irrespective of serotype, and with high uncertainty in Malawi due to small case numbers (Fig S4, Fig S5).

### Optimal age for vaccination

The optimal age for vaccination, the age of a single-dose vaccination that could prevent most IPD cases, is attained when vaccines are given earlier in the considered age range in the LIC/MIC setting, but not for England (Fig 2). In the base scenario with rapid waning of PPV23 VE, we found that the highest proportion of all IPD cases could be preventable if adults aged 55y are vaccinated in Brazil (30·5%, 22·4-43·3), in Malawi (31·0%, 7·7-96·6), and in South Africa (20·0%, 13·7-28·7), and if adults aged 70y are vaccinated in England (13·7%, 10·8-17·4) compared to a scenario without vaccination (Table S1). Also, In the base scenario with rapid waning VE, higher proportion of all preventable cases are estimated for using PCV20 vs PPV23 relative to a scenario without vaccination in Brazil (61·3%, 42·7-89·2) vs (30·1%, 20·6-43·5), Malawi (64·5 %, 13·0-92·2) vs (28·5%, 5·7-73·3), and South Africa (53·1%, 39·7-73·2) vs (19·8%, 13·0-29·2) among adults aged 55y, and in England (27·2%, 21·6-34·5) vs (13·6%, 10·4-17·6) among adults aged 70y, respectively (Table S2). Furthermore, higher proportion of IPD cases are preventable under slow vs rapid waning of PPV23 VE among adults aged 55y in Brazil (40·7%, 29·0-57·1) vs (30·1%, 20·6-43·5), Malawi (38·6 %, 8·0-77·2) vs (28·5%, 5·7-73·3), South Africa (26·4%, 18·2-38·3) vs (19·8%, 13·0-29·2), and among adults aged 70y in England (17·5%, 13·8-22·0) vs (13·6%, 10·4-17·6), respectively (Table S3).

**Figure 2.**
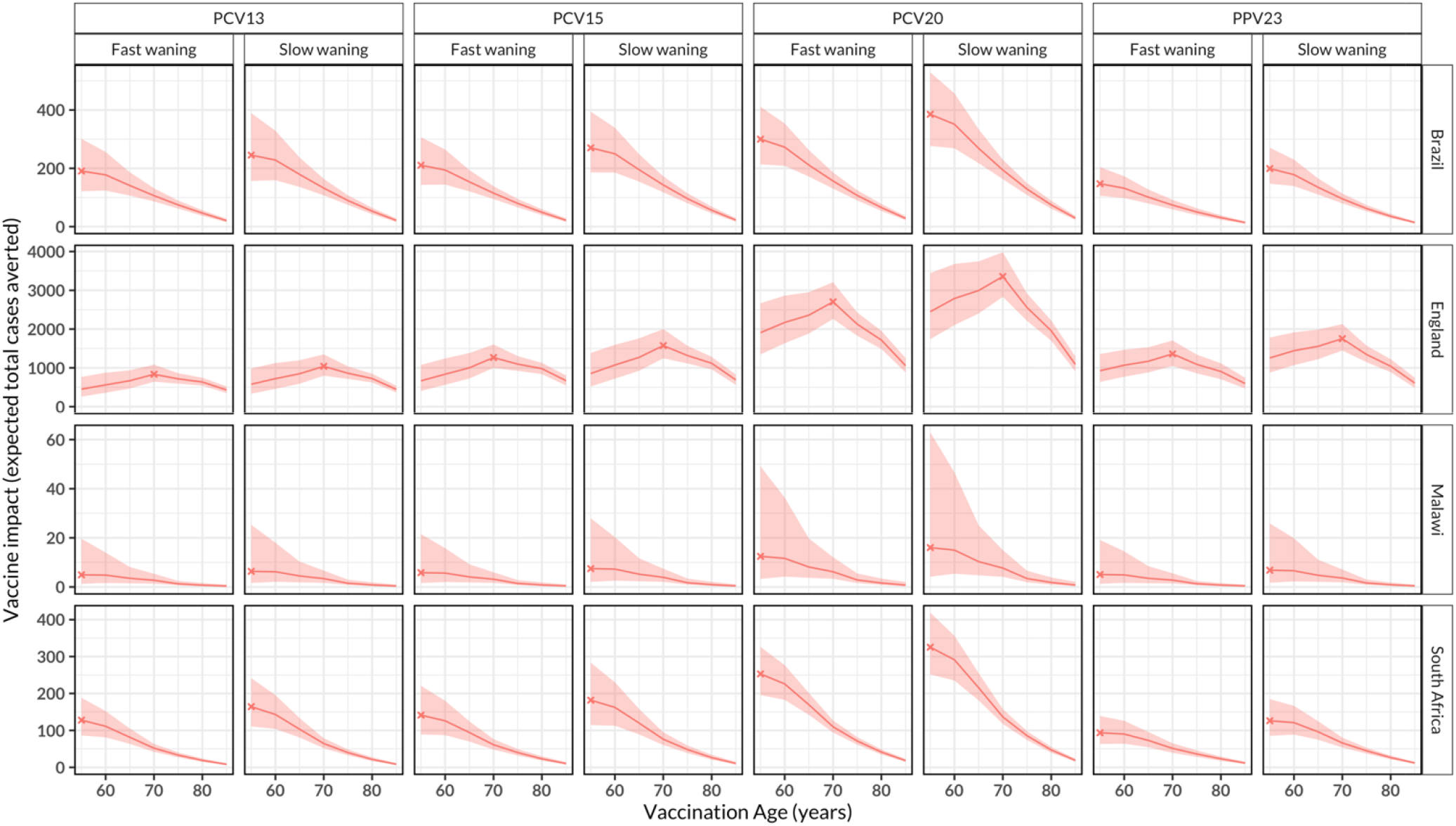
The impact of routine pneumococcal vaccination in older adults aged ≥55 years old (y). The expected absolute number of total IPD cases averted for the rest of age cohort lifetime by vaccinating every older adult in the age cohort stratified by country and vaccine product, under the scenario of age-independent initial vaccine efficacy/effectiveness (VE) and waning VE in Brazil, England, Malawi and South Africa. The lines represent cohort model mean estimates and the shaded ribbon represents 95% bootstrap confidence intervals for the mean estimates. The X corresponds to the optimal age for pneumococcal vaccination. In Brazil, Malawi and South Africa, most cases are preventable at age 55y whereas in England this is achieved at age 70y.

The optimal age for vaccinating the lowest number of individuals with single dose pneumococcal vaccine to prevent a reported case of IPD (vaccination efficiency) is estimated to differ by country, vaccine product or waning VE assumption (Fig 3). In a base case scenario with rapid waning of PPV23 VE, optimal age for vaccination efficiency is achieved through vaccination at 60y in Malawi 684 (244-2,009), 65y in Brazil 10,283 (8,165-13,162), and 75y in South Africa 3,643 (3,004-4,555) and at 80y in England 352 (287-438) (Table S4).

**Figure 3.**
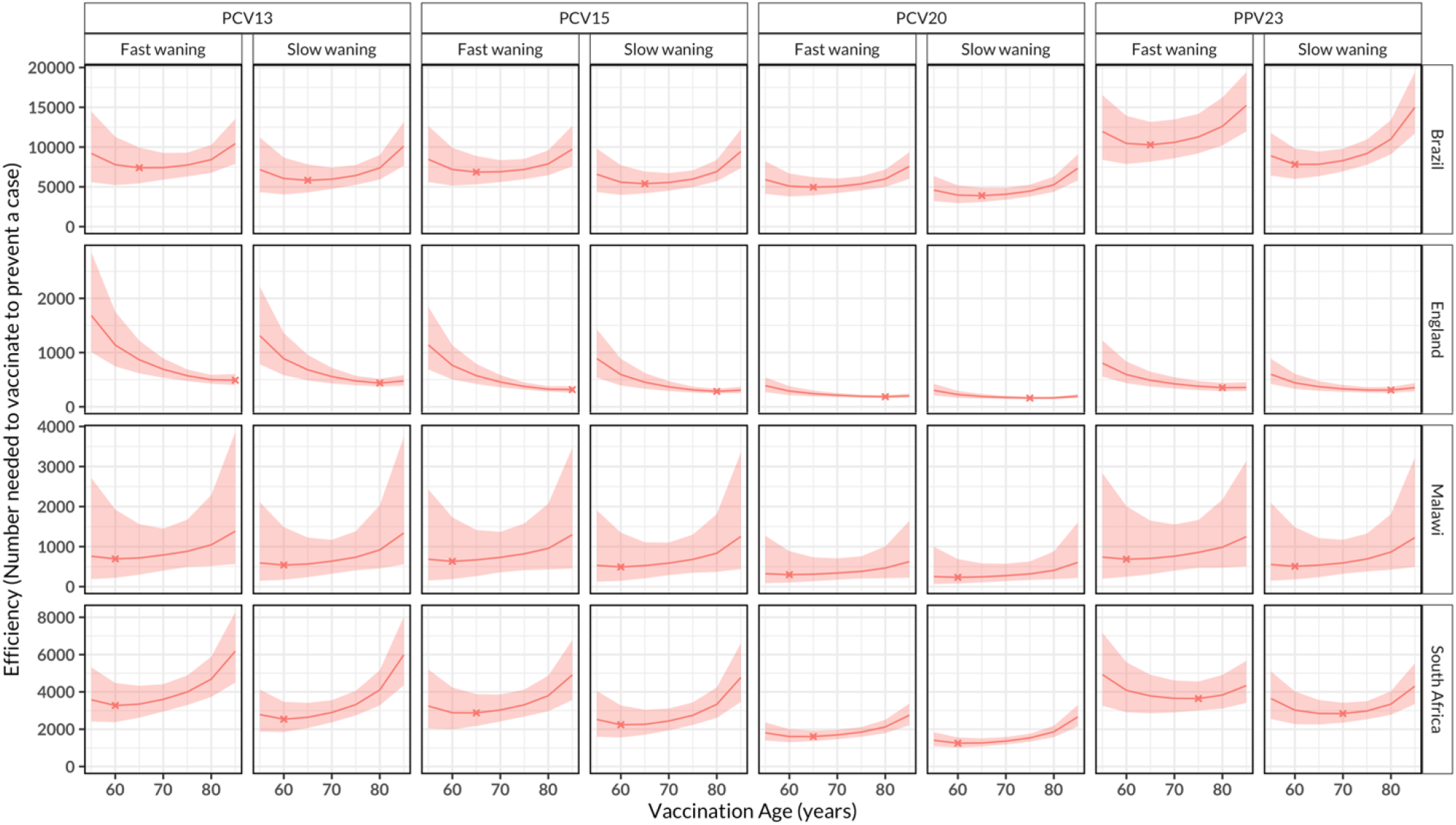
The efficiency of routine pneumococcal vaccination in older adults aged ≥55 years old (y). The number of individuals needed to vaccinate to prevent a case in each age of vaccination, stratified by country and vaccine product, under assumptions of age-independent initial vaccine efficacy/effectiveness (VE) and waning VE in Brazil, England, Malawi and South Africa. The lines represent cohort model mean estimates and the shaded ribbon represents 95% bootstrap confidence intervals for the mean estimates. The X represents the optimal age for efficiency of pneumococcal vaccination. Efficiency of vaccination varies by waning VE assumption and country reflecting sensitivity in reported invasive pneumococcal disease cases.

### Sensitivity analyses

If a scenario of age-dependent initial VE is considered in a sensitivity analysis, vaccine impact (total cases averted) remains maximum at 55y in Brazil, Malawi, and South Africa irrespective of vaccine product and assumption of waning VE as was the case with the base case scenario of age-independent initial VE. In contrast, optimal age for vaccination drops to 60y in England (Fig S6). In this scenario, the optimal age of vaccination efficiency is achieved by vaccination of 60y in Brazil, Malawi and South Africa, irrespective of vaccine product or assumptions of waning VE. In England, efficiency is achieved in 60y age cohort for PCV20 and PPV23 irrespective of waning VE assumption, and in 80y or 85y for PCV13’s and PCV15’s slow or fast waning VE, respectively (Fig S7).

## Discussion

We have assessed the optimal age-targeting for a single-dose PCV/PPV vaccination against VT-IPD in older adults ≥55y in Brazil, England, Malawi, and South Africa and find that vaccinating at 55y in considered LIC/MICs maximises preventable burden of IPD while vaccination at 70y is optimal in England. These findings suggest that the optimal age for vaccination may differ between countries and is driven by population age demographic, age-dependent IPD incidence and VE. Our findings were robust across pneumococcal vaccine products, and to alternative assumption on waning of VE.

The increasing incidence of IPD in older adulthood is outweighed by many more individuals in the fifties in LIC/MICs. The different age population structure in England, with a higher proportion of ≥55y in the middle to older adult ages, results in the optimal age for vaccination peaking later at 70y. The optimal age of vaccination in England also aligns with results from an Australian study that explored the role of timeliness in the cost-effectiveness of older adult vaccination and found that most deaths and hospitalisations were prevented if PCV13 was given to 70y, a peak age of prevented burden before dropping in remaining older ages [15].

The age with lowest number of individuals needed to vaccinate to prevent a case (optimal age of vaccination efficiency) differed by country, vaccine product or assumption of waning VE, in part reflecting the differences in sensitivity of IPD surveillance and the complex interplay between different factors influencing vaccination efficiency. Although this finding accounts for life expectancy, it does not consider quality-adjusted life-years losses due to IPD which could shift the optimal age of vaccination efficiency downwards because more individuals affected by IPD would be added to the fifties than elderly.

Our cohort model prediction of more preventable cases in all settings with the use of PCV20 vs PPV23 under similar conditions reflects higher efficacy/effectiveness of PCV20 than PPV23 [2]. Moreover, a larger number of IPD preventable cases by PCV20 than lesser-valent PCVs underlines the importance of additional serotypes 8, 10A, 11A, 12F and 15B and/or 22F, 33F covered by PCV20 in the remaining IPD burden in the mature infant-PCV era [10]. Generally, other disease endpoints including non-bacteraemic pneumonia may also be important for vaccine choice e.g., PCV13 is shown to be more efficacious in preventing VT non-bacteraemic pneumonia than PPV23 (46% vs 21%) [2], and if included in this analysis, it may likely heighten PCV13 impact to similar levels or higher than PPV23 in England, albeit with higher vaccine cost. With the likely upcoming use of PCV15 and PCV20 in infants in the near future [39], the vaccine preventable fraction of adult disease is likely to diminish correspondingly because of the indirect effects of such infant programme, and thus reducing the benefit of adult programmes that target the same serotypes as those targeted in infants. On the other hand, PCV21 targets most serotypes not targeted by PPV23 or PCV20 [40], which could likely improve tackling the remaining burden of pneumococcal disease in older adults, in the long absence of vaccines that target surface proteins common to all serotypes [41]. However, for LICs/MICs, PCV15, PCV20 and PCV21 may likely not be accessible soon due to their high initial costs.

Of note, we excluded older adults with HIV because they typically are considered separately for pneumococcal vaccination because of their high risk of disease before they reach old age. However, inclusion of older adults with HIV may likely elevate IPD burden in early older adulthood because of their huge presence in early than late adulthood due to survival bias thereby re-enforcing vaccinating early in older adulthood, particularly in Malawi and South Africa where adult HIV prevalence is relatively high [29]. In this analysis, the contribution of PCVs’ indirect protection from older adults to overall vaccine impact is ignored because older adults have low carriage rates unlike children who are the main transmitters, and the use of PPV23 does not protect against carriage and therefore unlikely to generate substantial herd immunity [42]. Sequential dosing of vaccine regimens was not modelled, although PCV15 followed by PPV23 is now recommended by the advisory committee on immunisation practices (ACIP) in the United States [43]. This sequential regimen would likely generate higher impact than a single dose of PCV20 or PPV23, albeit with cost-effectiveness implications that need further research, particularly in LICs [2].

PCVs and PPV23 are reported to have limited efficacy/effectiveness against serotype 3 pneumococcal carriage and disease [44], and our inclusion of serotype 3 IPD cases in this analysis imply that our vaccine impact estimates are likely biased upwards. PPV23 is already in use in ≥65y in England [17], and we show in this analysis that the effects of PPV23 use have reduced the original IPD burden (Fig 1), and without adjusting for the current PPV23 vaccination programme, our optimal age for vaccination in this country may be overestimated. Reported IPD case data in LMICs are usually incomplete or under-ascertained due to limited resources e.g. only 19 IPD cases were reported in Malawi and even less so when stratified by age and serotype [3]. Thus, our results should be cautiously interpreted. Despite potential biases of case underreporting, it seems reasonable to assume that underreporting is consistent across adult age groups, such that a relative change in IPD incidence by age to identify optimal age-targeting vaccination is less likely affected.

In conclusion, the optimal age-targeting for vaccination is largely driven by population age demographic, age-dependent IPD incidence and VE. In contrast to the typical use in adults in HICs, we find that pneumococcal vaccination in 55y older adults in LIC/MICs may be the most effective strategy in reducing the IPD burden among older adults without HIV, although affordability, cost-effectiveness and infant vaccination plans will also need to considered when designing a strategy to protect older adults against pneumococcal disease.

## Data Availability

An R script that was used to analyse the datasets and aggregated data are available in the GitHub repository.

https://github.com/deusthindwa/optimal.age.targeting.pneumo.vaccines

## Acknowledgements

We thank all the individuals working in respective invasive pneumococcal disease (IPD) surveillance and laboratory management teams in the four countries for providing important data on IPD isolates and related causing serotypes. DT, KCJ, SF NF, RSH, and TDS are supported by the National Institute for Health Research (NIHR) Global Health Research Unit on Mucosal Pathogens (MPRU) where RSH is a NIHR Senior Investigator. In addition, SF is supported by a Sir Henry Dale Fellowship jointly funded by the Wellcome Trust and the Royal Society. We would like to thank Stephen Gordon of Liverpool School of Tropical Medicine for insightful discussion and feedback at the initial stage of this work.

## Author contributions

Conceptualization; DT, SC, SF, EM, PM, NA.

Data curation; DT, JK, SM, TDS, ZAC, NFY, SCGA, MCdCB.

Formal analysis; DT, SC, SF.

Funding acquisition; DT, NF, SL, RSH, CC, MCdCB, SF.

Investigation; DT, SC, JK, AvG, SW, SM, TDS, EM, PM, NA, ZAC, NFY, KJ, NF, SCGA, SL, RSH, CC, MCdCB, SF.

Methodology; DT, SC, NF, SL, RSH, CC, MCdCB, SF

Project administration; JK, AvG, SW, SM, TDS, ZAC, NFY, KJ, NF, SCGA, SL, RSH, CC, MCdCB.

Resources; DT, NF, SL, RSH, CC, MCdCB, SF.

Software; DT, SC. Supervision; SF, NF

Validation; SC, JK, AvG, SW, SM, TDS, EM, PM, NA, ZAC, NFY, KJ, NF, SCGA, SL, RSH, CC, MCdCB, SF.

Visualization; DT, SC.

Writing - original draft; DT, SC, SF.

Writing - review & editing; DT, SC, JK, AvG, SW, SM, TDS, EM, PM, NA, ZAC, NFY, KJ, NF, SCGA, SL, RSH, CC, MCdCB, SF.

All authors read and approved the final manuscript.

## Data availability

An R script that was used to analyse the datasets and aggregated data are available in the GitHub repository https://github.com/deusthindwa/optimal.age.targeting.pneumo.vaccines.

## References

1. Wahl B, O’Brien KL, Greenbaum A, Majumder A, Liu L, Chu Y, et al. Burden of Streptococcus pneumoniae and Haemophilus influenzae type b disease in children in the era of conjugate vaccines: global, regional, and national estimates for 2000–15. Lancet Glob Health. 2018;6: e744–e757. doi:10.1016/S2214-109X(18)30247-X

2. Department of Immunization, Vaccines and Biologicals. WHO | SAGE Yellow Book for October 2020 [Internet]. Geneva, Switzerland: WHO; 2020 Oct p. 20. Available: http://www.who.int/immunization/sage/meetings/2020/october/presentations_background_docs/en/

3. Deloria Knoll M, Bennett JC, Garcia Quesada M, Kagucia EW, Peterson ME, Feikin DR, et al. Global Landscape Review of Serotype-Specific Invasive Pneumococcal Disease Surveillance among Countries Using PCV10/13: The Pneumococcal Serotype Replacement and Distribution Estimation (PSERENADE) Project. Microorganisms. Multidisciplinary Digital Publishing Institute; 2021;9: 742. doi:10.3390/microorganisms9040742

4. Bonnave C, Mertens D, Peetermans W, Cobbaert K, Ghesquiere B, Deschodt M, et al. Adult vaccination for pneumococcal disease: a comparison of the national guidelines in Europe. Eur J Clin Microbiol Infect Dis. 2019;38: 785–791. doi:10.1007/s10096-019-03485-3

5. Hurley D, Griffin C, Young M, Scott DA, Pride MW, Scully IL, et al. Safety, Tolerability, and Immunogenicity of a 20-Valent Pneumococcal Conjugate Vaccine (PCV20) in Adults 60 to 64 Years of Age. Clin Infect Dis. doi:10.1093/cid/ciaa1045

6. CDC Advisory Committee on Immunization Practices (ACIP). EtR for PCV15 use among adults ≥65 years old | CDC [Internet]. 27 Jan 2022 [cited 8 Feb 2022]. Available: https://www.cdc.gov/vaccines/acip/recs/grade/pneumo-PCV15-PPSV23-age-based-etr.html

7. Flasche S, Hoek AJV, Goldblatt D, Edmunds WJ, O’Brien KL, Scott JAG, et al. The Potential for Reducing the Number of Pneumococcal Conjugate Vaccine Doses While Sustaining Herd Immunity in High-Income Countries. PLOS Med. 2015;12: e1001839. doi:10.1371/journal.pmed.1001839

8. von Gottberg A, de Gouveia L, Tempia S, Quan V, Meiring S, von Mollendorf C, et al. Effects of Vaccination on Invasive Pneumococcal Disease in South Africa. N Engl J Med. 2014;371: 1889–1899. doi:10.1056/NEJMoa1401914

9. Bar-Zeev N, Swarthout TD, Everett DB, Alaerts M, Msefula J, Brown C, et al. Impact and effectiveness of 13-valent pneumococcal conjugate vaccine on population incidence of vaccine and non-vaccine serotype invasive pneumococcal disease in Blantyre, Malawi, 2006–18: prospective observational time-series and case-control studies. Lancet Glob Health. Elsevier; 2021;9: e989–e998. doi:10.1016/S2214-109X(21)00165-0

10. Garcia Quesada M, Yang Y, Bennett JC, Hayford K, Zeger SL, Feikin DR, et al. Serotype Distribution of Remaining Pneumococcal Meningitis in the Mature PCV10/13 Period: Findings from the PSERENADE Project. Microorganisms. Multidisciplinary Digital Publishing Institute; 2021;9: 738. doi:10.3390/microorganisms9040738

11. Swarthout TD, Fronterre C, Lourenço J, Obolski U, Gori A, Bar-Zeev N, et al. High residual carriage of vaccine-serotype Streptococcus pneumoniae after introduction of pneumococcal conjugate vaccine in Malawi. Nat Commun. Nature Publishing Group; 2020;11: 2222. doi:10.1038/s41467-020-15786-9

12. Thindwa D, Pinsent A, Ojal J, Gallagher KE, French N, Flasche S. Vaccine strategies to reduce the burden of pneumococcal disease in HIV-infected adults in Africa. Expert Rev Vaccines. Taylor & Francis; 2020;0: 1–8. doi:10.1080/14760584.2020.1843435

13. VACFA. Immunization Schedules - Africa | Vaccines for Africa [Internet]. 13 Apr 2018 [cited 27 Aug 2020]. Available: http://www.vacfa.uct.ac.za/immunization-schedules-africa

14. Matanock A. Use of 13-Valent Pneumococcal Conjugate Vaccine and 23-Valent Pneumococcal Polysaccharide Vaccine Among Adults Aged ≥65 Years: Updated Recommendations of the Advisory Committee on Immunization Practices. MMWR Morb Mortal Wkly Rep. 2019;68. doi:10.15585/mmwr.mm6846a5

15. Chen C, Wood JG, Beutels P, Menzies R, MacIntyre CR, Dirmesropian S, et al. The role of timeliness in the cost-effectiveness of older adult vaccination: A case study of pneumococcal conjugate vaccine in Australia. Vaccine. 2018;36: 1265–1271. doi:10.1016/j.vaccine.2018.01.052

16. Andrade AL, Minamisava R, Policena G, Cristo EB, Domingues CMS, Brandileone MC de C, et al. Evaluating the impact of PCV-10 on invasive pneumococcal disease in Brazil: A time-series analysis. Hum Vaccines Immunother. Taylor & Francis; 2016;12: 285–292. doi:10.1080/21645515.2015.1117713

17. Djennad A, Ramsay ME, Pebody R, Fry NK, Sheppard C, Ladhani SN, et al. Effectiveness of 23-Valent Polysaccharide Pneumococcal Vaccine and Changes in Invasive Pneumococcal Disease Incidence from 2000 to 2017 in Those Aged 65 and Over in England and Wales. EClinicalMedicine. 2019;6: 42–50. doi:10.1016/j.eclinm.2018.12.007

18. Cohen C, Mollendorf C von, Gouveia L de, Lengana S, Meiring S, Quan V, et al. Effectiveness of the 13-valent pneumococcal conjugate vaccine against invasive pneumococcal disease in South African children: a case-control study. Lancet Glob Health. 2017;5: e359–e369. doi:10.1016/S2214-109X(17)30043-8

19. Choi YH, Andrews N, Miller E. Estimated impact of revising the 13-valent pneumococcal conjugate vaccine schedule from 2+1 to 1+1 in England and Wales: A modelling study. PLOS Med. 2019;16: e1002845. doi:10.1371/journal.pmed.1002845

20. Brandileone M-CC, Almeida SCG, Minamisava R, Andrade A-L. Distribution of invasive Streptococcus pneumoniae serotypes before and 5 years after the introduction of 10-valent pneumococcal conjugate vaccine in Brazil. Vaccine. 2018;36: 2559–2566. doi:10.1016/j.vaccine.2018.04.010

21. Andrews NJ, Waight PA, George RC, Slack MPE, Miller E. Impact and effectiveness of 23-valent pneumococcal polysaccharide vaccine against invasive pneumococcal disease in the elderly in England and Wales. Vaccine. 2012;30: 6802–6808. doi:10.1016/j.vaccine.2012.09.019

22. Neto JT, Tannus Branco de Araujo G, Gagliardi A, Pinho A, Durand L, Fonseca M. Cost-effectiveness analysis of pneumococcal polysaccharide vaccination from age 60 in São Paulo State, Brazil. Hum Vaccin. Taylor & Francis; 2011;7: 1037–1047. doi:10.4161/hv.7.10.15987

23. Boyles TH, Brink A, Calligaro GL, Cohen C, Dheda K, Maartens G, et al. South African guideline for the management of community-acquired pneumonia in adults. J Thorac Dis. 2017;9: 1469–1502. doi:10.21037/jtd.2017.05.31

24. WasmÆlia B, Nuno Duarte da Costa B, Marcia Maria Melo Q, Wadih Joao Scandar N, Paulo CØsar Morae S, David Wu T, et al. Censo Demografico 2010 Brazil. Características da população e dos domicílios Resultados do universo [Internet]. Rio de Janeiro, RJ - Brasil: Instituto Brasileiro de Geografi a e Estatística; 2011 pp. 1–270. Report No.: 3. Available: https://biblioteca.ibge.gov.br/visualizacao/periodicos/93/cd_2010_caracteristicas_populacao_domicilios.pdf

25. Office for National Statistics UK. Estimates of the Population for the UK, England and Wales, Scotland and Northen Ireland 2017 [Internet]. Office for National Statistics; 2017 Jun. Available: https://www.ons.gov.uk/peoplepopulationandcommunity/populationandmigration/populationestimates

26. National Statistical Office Malawi. Malawi Population and Housing Census 2018 [Internet]. Zomba, Malawi, and Rockville, Maryland, USA: National Statistical Office Malawi; 2019 May pp. 1–311. Report No.: 4. Available: http://www.nsomalawi.mw/index.php%3Foption%3Dcom_content%26view%3Darticle%26id%3D226:2018-malawi-population-and-housing-census%26catid%E2%80%89%3D%E2%80%898:reports%26Itemid%E2%80%89%3D%E2%80%896

27. Statistics South Africa. South Africa Population Census 2011 [Internet]. Statistics South Africa; 2011. Available: http://www.statssa.gov.za/?page_id=3992

28. World Bank Group. World Population Annual Growth [Internet]. 10 Aug 2021 [cited 10 Aug 2021]. Available: https://data.worldbank.org/indicator/SP.POP.GROW?

29. Johnson L, Dorrington R. Thembisa Project - A Mathematical Model of South African HIV epidemic [Internet]. 8 Feb 2022 [cited 8 Feb 2022]. Available: https://www.thembisa.org/

30. Fox GA, Kendall BE. Demographic Stochasticity and the Variance Reduction Effect. Ecology. 2002;83: 1928–1934. doi:10.1890/0012-9658(2002)083[1928:DSATVR]2.0.CO;2

31. Chaguza C, Heinsbroek E, Gladstone RA, Tafatatha T, Alaerts M, Peno C, et al. Early Signals of Vaccine-driven Perturbation Seen in Pneumococcal Carriage Population Genomic Data. Clin Infect Dis. 2020;70: 1294–1303. doi:10.1093/cid/ciz404

32. Dwyer-Lindgren L, Cork MA, Sligar A, Steuben KM, Wilson KF, Provost NR, et al. Mapping HIV prevalence in sub-Saharan Africa between 2000 and 2017. Nature. 2019;570: 189. doi:10.1038/s41586-019-1200-9

33. Wright LB, Hughes GJ, Chapman KE, Gorton R, Wilson D. Effectiveness of the 23-valent pneumococcal polysaccharide vaccine against invasive pneumococcal disease in people aged 65 years and over in the North East of England, April 2006–July 2012. Trials Vaccinol. 2013;2: 45–48. doi:10.1016/j.trivac.2013.09.004

34. Rudnick W, Liu Z, Shigayeva A, Low DE, Green K, Plevneshi A, et al. Pneumococcal vaccination programs and the burden of invasive pneumococcal disease in Ontario, Canada, 1995–2011. Vaccine. 2013;31: 5863–5871. doi:10.1016/j.vaccine.2013.09.049

35. Rodríguez MAG, Gavín MAO, García-Comas L, Moreno JCS, Deorador EC, Carbajo MDL, et al. Effectiveness of 23-valent pneumococcal polysaccharide vaccine in adults aged 60 years and over in the Region of Madrid, Spain, 2008–2011. Eurosurveillance. European Centre for Disease Prevention and Control; 2014;19: 20922. doi:10.2807/1560-7917.ES2014.19.40.20922

36. Bonten MJM, Huijts SM, Bolkenbaas M, Webber C, Patterson S, Gault S, et al. Polysaccharide Conjugate Vaccine against Pneumococcal Pneumonia in Adults. N Engl J Med. 2015;372: 1114–1125. doi:10.1056/NEJMoa1408544

37. Patterson S, Webber C, Patton M, Drews W, Huijts SM, Bolkenbaas M, et al. A post hoc assessment of duration of protection in CAPiTA (Community Acquired Pneumonia immunization Trial in Adults). Trials Vaccinol. 2016;5: 92–96. doi:10.1016/j.trivac.2016.04.004

38. van Werkhoven CH, Huijts SM, Bolkenbaas M, Grobbee DE, Bonten MJM. The Impact of Age on the Efficacy of 13-valent Pneumococcal Conjugate Vaccine in Elderly. Clin Infect Dis. 2015;61: 1835–1838. doi:10.1093/cid/civ686

39. Senders S, Klein NP, Lamberth E, Thompson A, Drozd J, Trammel J, et al. Safety and Immunogenicity of a 20-valent Pneumococcal Conjugate Vaccine in Healthy Infants in the United States. Pediatr Infect Dis J. 2021;40: 944–951. doi:10.1097/INF.0000000000003277

40. Merck. Merck Announces U.S. FDA has Granted Breakthrough Therapy Designation for V116, the Company’s Investigational 21-Valent Pneumococcal Conjugate Vaccine, for the Prevention of Invasive Pneumococcal Disease and Pneumococcal Pneumonia in Adults. In: Merck.com [Internet]. [cited 10 Jul 2022]. Available: https://www.merck.com/news/merck-announces-u-s-fda-has-granted-breakthrough-therapy-designation-for-v116-the-companys-investigational-21-valent-pneumococcal-conjugate-vaccine-for-the-prevention-of-invasive-pneumococ/

41. Weinberger DM, Harboe ZB, Shapiro ED. Developing Better Pneumococcal Vaccines for Adults. JAMA Intern Med. 2017;177: 303–304. doi:10.1001/jamainternmed.2016.8289

42. Flasche S, Lipsitch M, Ojal J, Pinsent A. Estimating the contribution of different age strata to vaccine serotype pneumococcal transmission in the pre vaccine era: a modelling study. BMC Med. 2020;18: 129. doi:10.1186/s12916-020-01601-1

43. Kobayashi M. Use of 15-Valent Pneumococcal Conjugate Vaccine and 20-Valent Pneumococcal Conjugate Vaccine Among U.S. Adults: Updated Recommendations of the Advisory Committee on Immunization Practices — United States, 2022. MMWR Morb Mortal Wkly Rep. 2022;71. doi:10.15585/mmwr.mm7104a1

44. Linley E, Bell A, Gritzfeld JF, Borrow R. Should Pneumococcal Serotype 3 Be Included in Serotype-Specific Immunoassays? Vaccines. Multidisciplinary Digital Publishing Institute; 2019;7: 4. doi:10.3390/vaccines7010004

