## Supplementary for "Optimal age targeting for pneumococcal vaccination in older adults; a modelling study"

**Table of Contents**

Text S1- Description of IPD surveillances in Brazil, England, Malawi and South Africa

Text S2 - Description of the back-inflation method to correct for original IPD cases in England

Text S3 - Description of the exponential growth model fitted to IPD cases data

Table S1 - Comparing preventable IPD cases between age cohorts

Table S2 - Comparing preventable IPD cases under the use of different vaccine products

Table S3 - Comparing preventable IPD cases between fast and slow efficacy waning

Table S4 - Comparing numbers needed to vaccinate to prevent a single case (vaccination efficiency)

Figure S1 - IPD cases in annual age stratified by serotype group, year of surveillance and country

Figure S2 - Reported initial efficacy/effectiveness (VE) and waning VE of pneumococcal vaccines

Figure S3 - Estimated initial VE and waning VE from time since vaccination

Figure S4 - Expected burden of IPD cases across age stratified by serotype group and country

Figure S5 - Age-scaled IPD incidence in age groups stratified by serotype group and country

Figure S6 - The impact of routine pneumococcal vaccination in older adults under the age-dependent initial VE

Figure S7 - The efficiency of routine pneumococcal vaccination in older adults under the age-dependent initial VE

**Text S1: Invasive pneumococcal disease surveillance**

In Brazil, England, Malawi and South Africa, invasive pneumococcal disease (IPD) in adults is part of their national or subnational disease surveillance. In Brazil, there is population-based surveillance only for meningitis, which are notifiable diseases. For IPD, there is laboratory-based surveillance. The invasive isolates isolated all over the country are sent to the Centre of Bacteriology at Adolfo Lutz Institute (IAL), the Brazilian National Reference Laboratory, for meningitis and IPD characterization. IAL receives IPD isolates collected from private hospitals and laboratories, and from the national network of 25 laboratories coordinated by Brazilian Ministry of Health, with each laboratory covering each Brazilian state [1]. In England, the National Health Service (NHS) hospital laboratories routinely submit IPD isolates to the UK Health Security Agency (UKHSA) for confirmation and serotyping with corresponding electronic reports sent via second generation surveillance system (SGSS) [2]. Reports without isolates are followed-up by UKHSA to request isolate referral to ensure consistently high serotyping rates. In Malawi, an ongoing sentinel surveillance for the laboratory confirmed IPD at a government referral hospital, Queen Elizabeth Central Hospital (QECH), in Blantyre has been in place since 1998, serving now 1.3 million residents of Blantyre district. IPD isolates are submitted to the Malawi Liverpool Wellcome Clinical Research Programme laboratory sitting next to QECH through surveillance programme of IN-patients and Epidemiology system where confirmation and serotyping are conducted [3,4]. In South Africa, the Group for Enteric, Respiratory and Meningeal disease Surveillance in South Africa (GERMS-SA) conducts national, active laboratory-based surveillance across South Africa in a network consisting of nearly 130 public and private microbiology laboratories. Each laboratory submits pneumococcal isolates a long with patient demographics to the Centre for Respiratory Diseases and Meningitis (CRDM) at the National Institute for Communicable Diseases where confirmation and serotyping are performed [5,6].

**Text S2: Back-inflation method to correct for IPD cases in England due to the presence of PPV23 vaccination**

To estimate the number of English IPD cases assuming absence of PPV23 vaccination in the population, we combined data on PPV23 vaccination coverage (70%), observed number of IPD cases in the presence of PPV23 vaccination program, and age-adjusted vaccine effectiveness of PPV23 of 41% from 0 to <2 years, 34% 2 to <5 years, and 23% from 5+ years as estimated by Djennad et al. The formula below was used to estimate the new number of IPD cases (inflated) assuming the absence of PPV23 vaccination in the older English adult population.

$$C_{I}=\frac{C_{o}}{\nu*\left( 1-{VE}_{a} \right)+(1-v)}\ldots\ldots\ldots\ldots\ldots\ldots\ldots\ldots\ldots\ldots\ldots\ldots(1)$$

Where $C_{I}$ is the total IPD cases assuming no PPV23 vaccination in the older English population, $C_{o}$ is the observed total IPD cases in the presence of PPV23 vaccination program, $\nu$ is the PPV23 coverage, and ${VE}_{a}$ is the age ($a$) adjusted PPV23 vaccine effectiveness from time since vaccination as estimated by Djennad et al.

**Text S3: Exponential growth model**

To interpolate and extrapolate IPD incidence to annual ages from 55 to 90 years old adults, a nonlinear model (NLS) was fitted to the reported age-specific IPD incidences, minimising the least-squares difference between the model and data using Gauss-Newton algorithm (NLS) as depicted in the main analysis [7]. Initial values of the intercept ($\alpha$) and gradient ($\beta$) for the NLS model were obtained from a fitted linear model, as shown in equation 1.

$$Log\left( I|\alpha\right)=Log\left( \gamma\right)+\beta*\alpha\ldots\ldots\ldots\ldots\ldots\ldots\ldots\ldots\ldots\ldots\ldots\ldots\ldots(2)$$

Where $I$ is the IPD incidence for each age group $\alpha$, $\gamma$ is the model intercept, and $\beta$ is the model gradient (growth rate) needed as initial values for equation 2.

| **Table S1**. A base case scenario, age-independent initial vaccine efficacy/effectiveness (VE), using PPV23 under a fast waning VE. Reduction in vaccine-type IPD cases in different vaccination age cohorts (e.g., 55 vs 65 years cohorts) relative to no vaccination scenario. | | | | |
| --- | --- | --- | --- | --- |
| **Country** | **Vaccination age** | **2.50%** | **50%** | **97.50%** |
| Brazil | 55 | 22.4 | 30.5 | 43.3 |
| Brazil | 60 | 20.9 | 27.3 | 36.8 |
| Brazil | 65 | 16.8 | 20.9 | 26.8 |
| Brazil | 70 | 12.3 | 15.3 | 18.9 |
| Brazil | 75 | 8.4 | 10.4 | 12.9 |
| Brazil | 80 | 4.9 | 6.4 | 8.0 |
| Brazil | 85 | 2.2 | 2.9 | 3.8 |
| England | 55 | 6.3 | 9.2 | 13.9 |
| England | 60 | 7.7 | 10.6 | 15.0 |
| England | 65 | 8.9 | 11.7 | 15.6 |
| England | 70 | 10.8 | 13.7 | 17.4 |
| England | 75 | 8.7 | 11.0 | 13.5 |
| England | 80 | 7.3 | 9.1 | 11.1 |
| England | 85 | 4.8 | 6.0 | 7.5 |
| Malawi | 55 | 7.7 | 31.0 | 96.6 |
| Malawi | 60 | 9.6 | 30.5 | 85.9 |
| Malawi | 65 | 9.0 | 22.4 | 49.3 |
| Malawi | 70 | 8.2 | 17.3 | 35.2 |
| Malawi | 75 | 4.0 | 7.9 | 16.2 |
| Malawi | 80 | 2.0 | 4.6 | 10.4 |
| Malawi | 85 | 0.9 | 2.4 | 6.5 |
| South Africa | 55 | 13.7 | 20.0 | 28.7 |
| South Africa | 60 | 13.8 | 19.2 | 25.9 |
| South Africa | 65 | 11.9 | 15.4 | 20.0 |
| South Africa | 70 | 8.5 | 10.8 | 13.7 |
| South Africa | 75 | 6.1 | 7.7 | 9.6 |
| South Africa | 80 | 3.8 | 4.9 | 6.1 |
| South Africa | 85 | 1.9 | 2.5 | 3.2 |

| **Table S2**. A base case scenario, age-independent initial efficacy/effectiveness (VE), where 55- and 70-years old cohorts are vaccinated under fast waning VE. Reduction in vaccine-type IPD cases between use of different vaccines products (e.g., PCV20 vs PPV23) relative to no vaccination scenario. | | | | | |
| --- | --- | --- | --- | --- | --- |
| **Country** | **Vaccination age** | **Serotype group** | **2.50%** | **50%** | **97.50%** |
| Brazil | 55 | PCV13 | 24.7 | 38.4 | 61.5 |
| Brazil | 70 | PCV13 | 16.5 | 21.7 | 28.7 |
| Brazil | 55 | PCV15 | 27.8 | 43.8 | 65.0 |
| Brazil | 70 | PCV15 | 18.4 | 23.5 | 30.3 |
| Brazil | 55 | PCV20 | 42.7 | 61.3 | 89.2 |
| Brazil | 70 | PCV20 | 25.1 | 32.0 | 40.4 |
| Brazil | 55 | PPV23 | 20.6 | 30.1 | 43.5 |
| Brazil | 70 | PPV23 | 11.2 | 15.2 | 20.0 |
| England | 55 | PCV13 | 2.5 | 4.4 | 7.7 |
| England | 70 | PCV13 | 6.2 | 8.3 | 11.1 |
| England | 55 | PCV15 | 3.9 | 6.6 | 11.0 |
| England | 70 | PCV15 | 9.5 | 12.6 | 17.0 |
| England | 55 | PCV20 | 13.3 | 19.2 | 29.2 |
| England | 70 | PCV20 | 21.6 | 27.2 | 34.5 |
| England | 55 | PPV23 | 6.2 | 9.3 | 13.7 |
| England | 70 | PPV23 | 10.4 | 13.6 | 17.6 |
| Malawi | 55 | PCV13 | 5.7 | 30.9 | 74.0 |
| Malawi | 70 | PCV13 | 5.5 | 16.1 | 46.0 |
| Malawi | 55 | PCV15 | 6.4 | 32.2 | 75.3 |
| Malawi | 70 | PCV15 | 6.0 | 17.9 | 50.0 |
| Malawi | 55 | PCV20 | 13.0 | 64.5 | 92.2 |
| Malawi | 70 | PCV20 | 11.7 | 37.3 | 99.0 |
| Malawi | 55 | PPV23 | 5.7 | 28.5 | 73.3 |
| Malawi | 70 | PPV23 | 5.5 | 15.8 | 43.1 |
| South Africa | 55 | PCV13 | 17.8 | 26.7 | 40.8 |
| South Africa | 70 | PCV13 | 8.6 | 11.0 | 13.9 |
| South Africa | 55 | PCV15 | 18.6 | 29.7 | 48.7 |
| South Africa | 70 | PCV15 | 10.0 | 12.9 | 17.2 |
| South Africa | 55 | PCV20 | 39.7 | 53.1 | 73.2 |
| South Africa | 70 | PCV20 | 18.9 | 23.2 | 28.4 |
| South Africa | 55 | PPV23 | 13.0 | 19.8 | 29.2 |
| South Africa | 70 | PPV23 | 8.1 | 10.8 | 13.8 |

| **Table S3**. A base case scenario, age-independent initial efficacy/effectiveness (VE), of PPV23 use in 65 years old cohort. Reduction in vaccine-type IPD cases between fast vs slow waning VE relative to no vaccination scenario. | | | | | | |
| --- | --- | --- | --- | --- | --- | --- |
| **Country** | **Serotype group** | **Waning VE** | **Vaccination age** | **2.50%** | **50%** | **97.50%** |
| Brazil | PCV20 | Fast waning | 55 | 42.7 | 61.3 | 89.2 |
| Brazil | PCV20 | Slow waning | 55 | 55.0 | 79.0 | 94.3 |
| Brazil | PPV23 | Fast waning | 55 | 20.6 | 30.1 | 43.5 |
| Brazil | PPV23 | Slow waning | 55 | 29.0 | 40.7 | 57.1 |
| England | PCV20 | Fast waning | 55 | 13.3 | 19.2 | 29.2 |
| England | PCV20 | Slow waning | 55 | 17.1 | 24.8 | 37.4 |
| England | PPV23 | Fast waning | 55 | 6.2 | 9.3 | 13.7 |
| England | PPV23 | Slow waning | 55 | 8.3 | 12.4 | 18.6 |
| Malawi | PCV20 | Fast waning | 55 | 13.0 | 64.5 | 83.2 |
| Malawi | PCV20 | Slow waning | 55 | 16.7 | 83.0 | 96.6 |
| Malawi | PPV23 | Fast waning | 55 | 5.7 | 28.5 | 73.3 |
| Malawi | PPV23 | Slow waning | 55 | 8.0 | 38.6 | 77.2 |
| South Africa | PCV20 | Fast waning | 55 | 39.7 | 53.1 | 73.2 |
| South Africa | PCV20 | Slow waning | 55 | 51.2 | 68.2 | 93.8 |
| South Africa | PPV23 | Fast waning | 55 | 13.0 | 19.8 | 29.2 |
| South Africa | PPV23 | Slow waning | 55 | 18.2 | 26.4 | 38.3 |
| Brazil | PCV20 | Fast waning | 70 | 25.1 | 32.0 | 40.4 |
| Brazil | PCV20 | Slow waning | 70 | 31.2 | 39.8 | 50.2 |
| Brazil | PPV23 | Fast waning | 70 | 11.2 | 15.2 | 20.0 |
| Brazil | PPV23 | Slow waning | 70 | 15.3 | 19.5 | 25.0 |
| England | PCV20 | Fast waning | 70 | 21.6 | 27.2 | 34.5 |
| England | PCV20 | Slow waning | 70 | 27.0 | 33.9 | 42.5 |
| England | PPV23 | Fast waning | 70 | 10.4 | 13.6 | 17.6 |
| England | PPV23 | Slow waning | 70 | 13.8 | 17.5 | 22.0 |
| Malawi | PCV20 | Fast waning | 70 | 11.7 | 37.3 | 99.0 |
| Malawi | PCV20 | Slow waning | 70 | 14.5 | 46.4 | 72.7 |
| Malawi | PPV23 | Fast waning | 70 | 5.5 | 15.8 | 43.1 |
| Malawi | PPV23 | Slow waning | 70 | 7.5 | 20.7 | 54.6 |
| South Africa | PCV20 | Fast waning | 70 | 18.9 | 23.2 | 28.4 |
| South Africa | PCV20 | Slow waning | 70 | 23.4 | 28.8 | 35.4 |
| South Africa | PPV23 | Fast waning | 70 | 8.1 | 10.8 | 13.8 |
| South Africa | PPV23 | Slow waning | 70 | 11.0 | 13.8 | 17.3 |

| **Table S4.** A base case scenario, age-independent initial vaccine efficacy/effectiveness (VE), of PPV23 use under fast waning VE. Number needed to vaccinate (NNV) to prevent a single IPD case. | | | | |
| --- | --- | --- | --- | --- |
| **Country** | **Vaccination age** | **2.50%** | **50%** | **97.50%** |
| Brazil | 55 | 8399 | 11947 | 16530 |
| Brazil | 60 | 7868 | 10461 | 13934 |
| Brazil | 65 | 8165 | 10283 | 13162 |
| Brazil | 70 | 8593 | 10592 | 13472 |
| Brazil | 75 | 9210 | 11271 | 14171 |
| Brazil | 80 | 10208 | 12602 | 16234 |
| Brazil | 85 | 11969 | 15238 | 19419 |
| England | 55 | 556 | 805 | 1222 |
| England | 60 | 434 | 594 | 834 |
| England | 65 | 374 | 487 | 647 |
| England | 70 | 336 | 423 | 544 |
| England | 75 | 307 | 378 | 475 |
| England | 80 | 287 | 352 | 438 |
| England | 85 | 292 | 355 | 450 |
| Malawi | 55 | 201 | 738 | 2835 |
| Malawi | 60 | 244 | 684 | 2009 |
| Malawi | 65 | 309 | 704 | 1649 |
| Malawi | 70 | 399 | 759 | 1552 |
| Malawi | 75 | 469 | 854 | 1663 |
| Malawi | 80 | 473 | 984 | 2171 |
| Malawi | 85 | 504 | 1248 | 3139 |
| South Africa | 55 | 3260 | 4931 | 7171 |
| South Africa | 60 | 2909 | 4083 | 5589 |
| South Africa | 65 | 2853 | 3783 | 4919 |
| South Africa | 70 | 2900 | 3652 | 4611 |
| South Africa | 75 | 3004 | 3643 | 4555 |
| South Africa | 80 | 3108 | 3830 | 4908 |
| South Africa | 85 | 3413 | 4338 | 5664 |

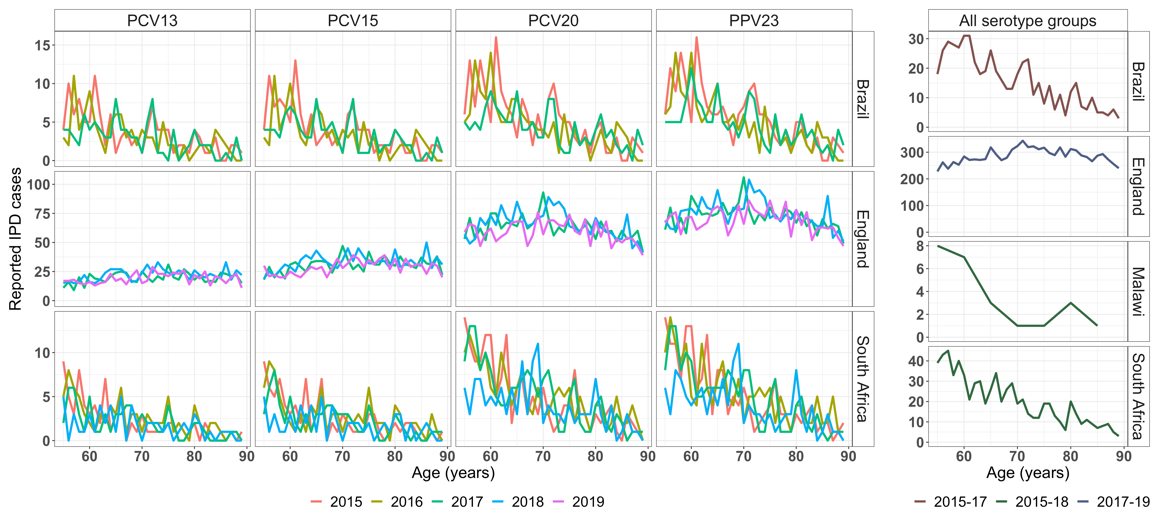

**Figure S1**. Distribution of invasive pneumococcal disease (IPD) cases in annual age, stratified by serotype group, year of surveillance and country. Serotype group specific IPD cases in Brazil (from 2015-2017), England (from 2017-2019) and South Africa (from 2015-2018) in annual age in older adults. Serogroup-specific IPD cases in each setting were average for all reported years and used to compute serotype group- and age-specific IPD incidence. Due to small sample size of IPD cases reported and incomplete serotyping data for age-group specific serotype distributions in Malawi, we calculated all-age serotype distribution and assumed the same in each age group.

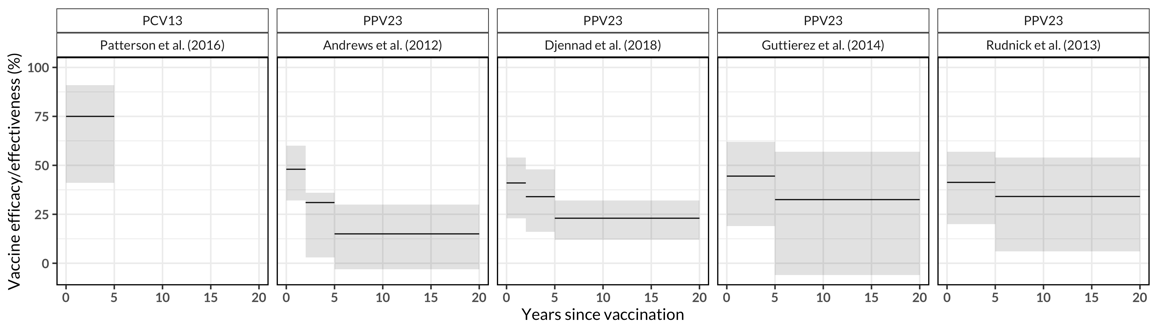

**Figure S2**. Reported initial vaccine efficacy/effectiveness (VE) and waning VE of pneumococcal polysaccharide vaccine (PPV23) and pneumococcal conjugate vaccine (PCV13), with bootstrapped 95% confidence intervals (line and ribbon) estimated from a piecewise constant model, with time since vaccination and stratification by study’s first author name. Initial VE and waning VE from Andrews et al. and Djennad et al. are used for the PPV23 in the vaccination impact cohort model as they provide the highest and lowest initial VE levels as well as fast and slow waning VE, respectively, whereas the initial VE from Patterson at al. (stable for 5 years) and fast waning VE from Andrews et al. and slow waning VE from Djennad et al. are used for all the PCVs (PCV13, PCV15, and PCV20).

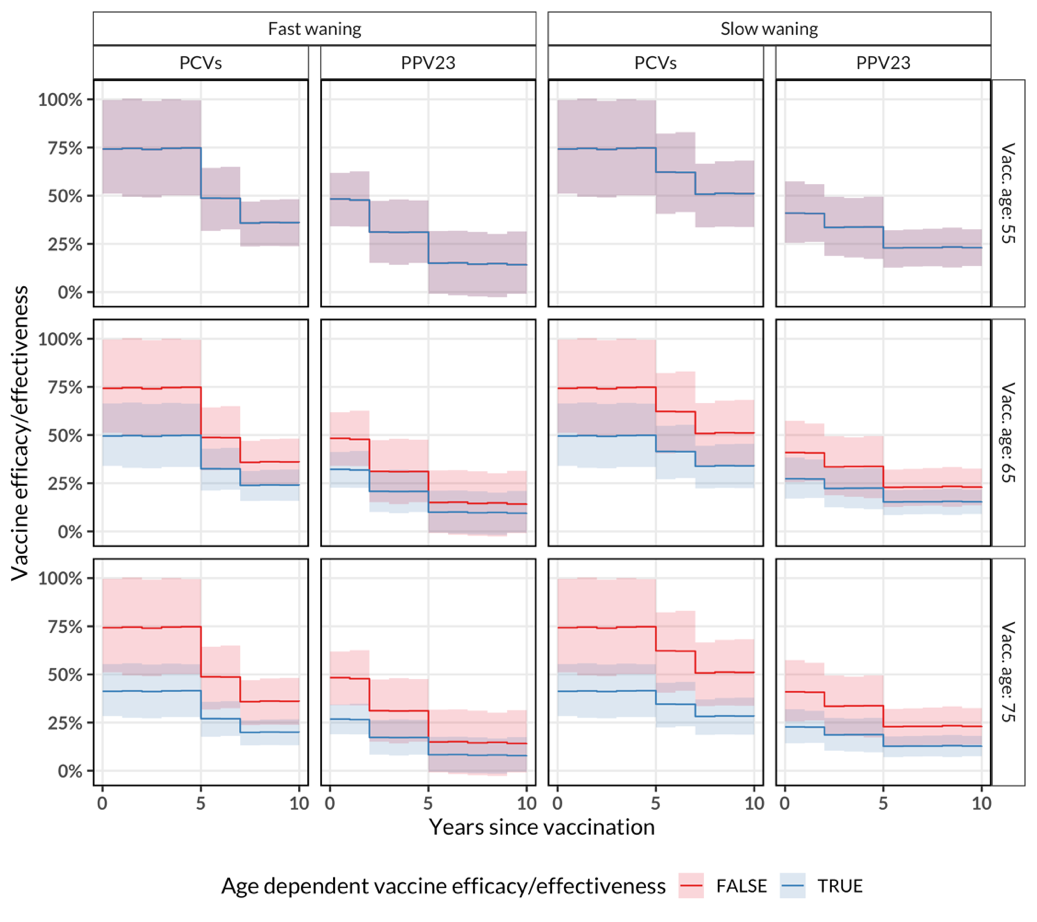

**Figure S3**. Reported initial vaccine efficacy/effectiveness (VE) and estimated waning VE of pneumococcal polysaccharide vaccine (PPV23) and pneumococcal conjugate vaccine (PCV13) with bootstrapped 95% confidence intervals (line and ribbon), estimated from a piecewise constant model, with time since vaccination and stratification by study’s first author name and *a priori* age of 65 and 75 years old at which initial VE is lower than 55 years old.

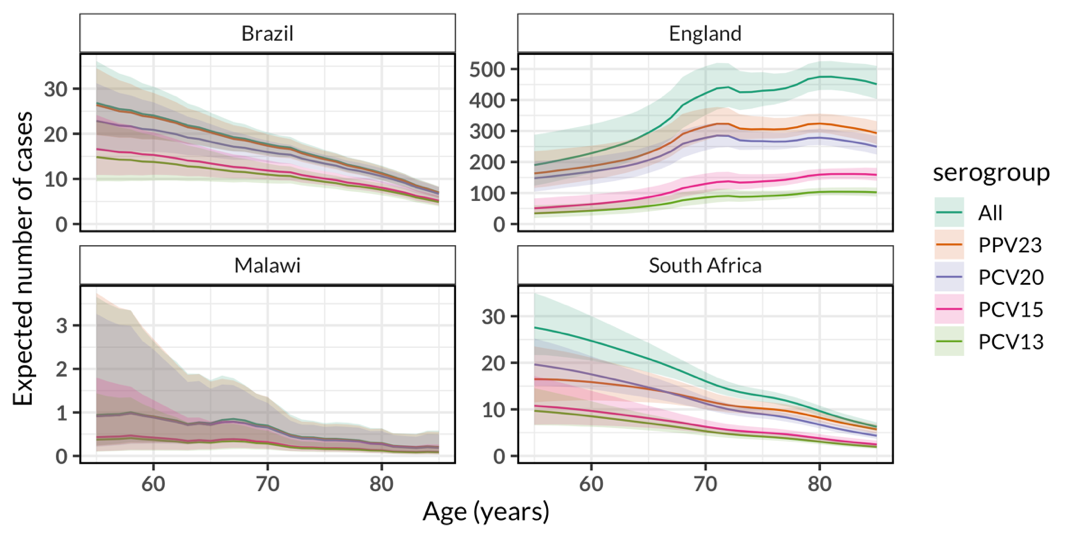

**Figure S4**. The expected burden of invasive pneumococcal disease (IPD) cases across age stratified by serotype group in Brazil (from 2015-2017), England (from 2017-2019) and South Africa (from 2015-2018) in annual age in older adults. Serotype group specific expected number of IPD cases in each setting was obtained by computing the product of observed IPD cases aggregated for all reported years and the fitted IPD incidence. Uncertainty in the expected number of cases was based on uncertainty in the fitted incidence obtained by bootstrap sampling 1000 times using the fitted parameter means and covariance matrix of a fitted exponential model.

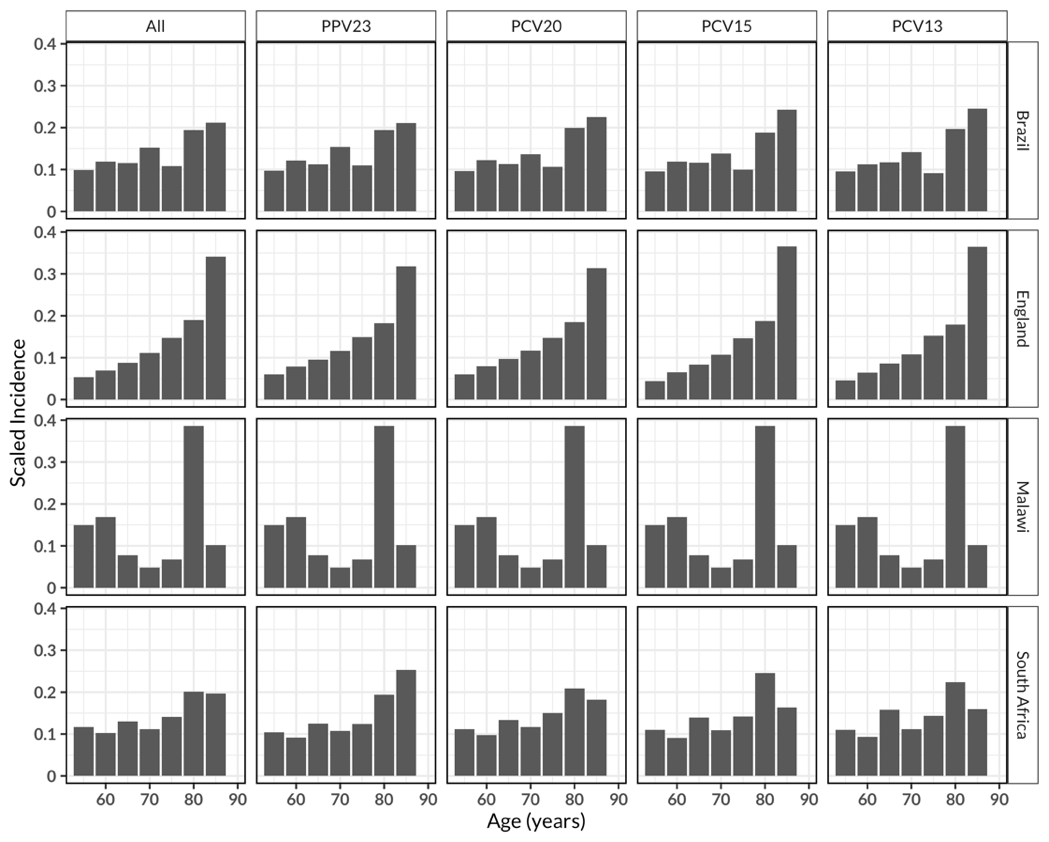

**Figure S5**. Age-scaled invasive pneumococcal disease (IPD) incidence in age groups stratified by serotype group and country for all years of IPD surveillance in a mature infant PCV era (at least four years post-PCV introduction). Age-scaled IPD incidence is estimated by dividing age group-specific IPD incidence by total incidence across all age groups within that serotype group. Age-scaled IPD incidence increases monotonically with increasing age irrespective of serotype group or country except for Malawi where the unstable signal in age-scaled IPD incidence reflects small numbers of reported IPD cases.

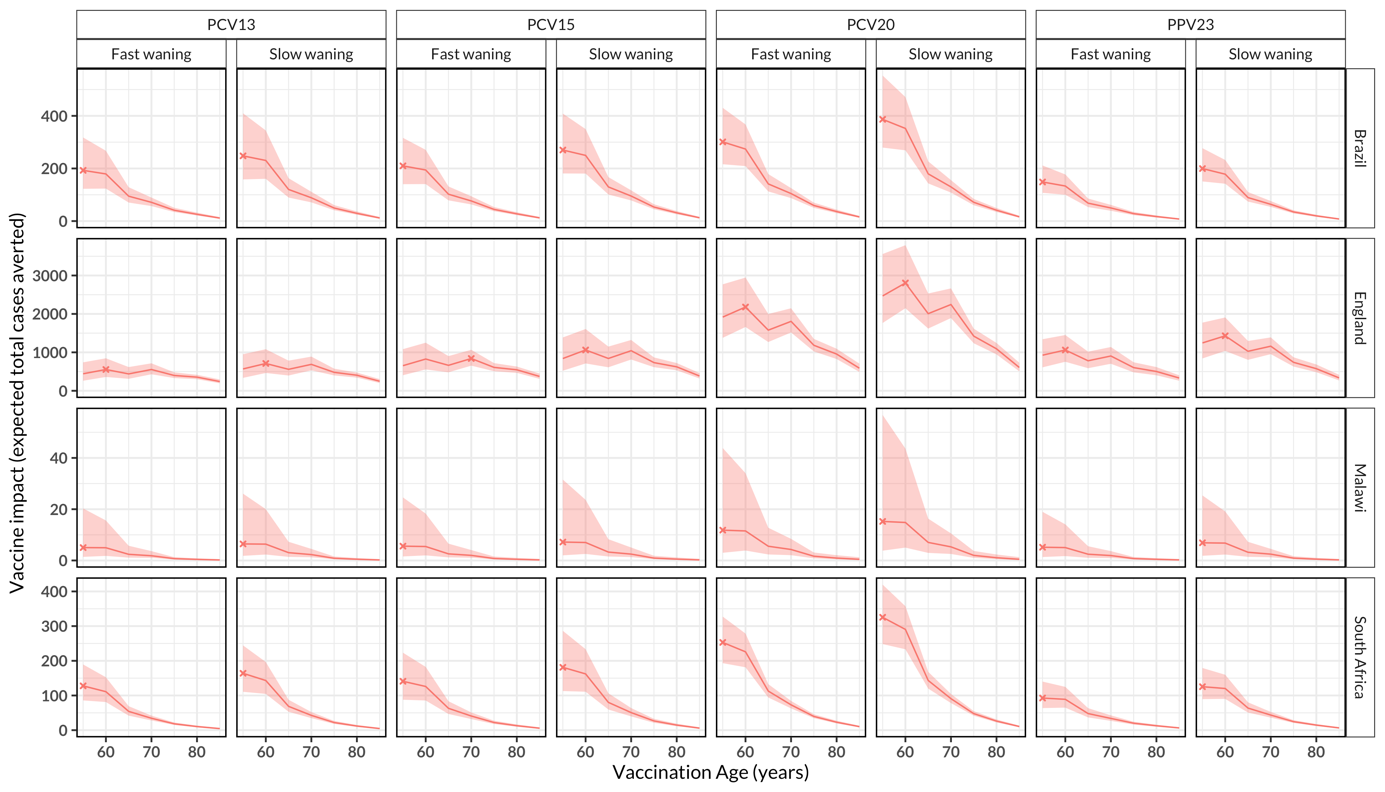

**Figure S6**. The impact of routine pneumococcal vaccination in older adults aged ≥55 years old (y). The expected absolute number of total IPD cases averted for the rest of age cohort lifetime by vaccinating every older adult in the age cohort stratified by country and vaccine product, under alternative scenario of of age-dependent initial vaccine efficacy/effectiveness (VE) and waning VE in Brazil, England, Malawi and South Africa. The lines represent cohort model mean estimates and the shaded ribbon represents 95% bootstrap confidence intervals for the mean estimates. The X represents the optimal age for pneumococcal vaccination. In Brazil, Malawi and South Africa, most cases are prevented at age 55y whereas in England this is achieved mostly at age 60y.

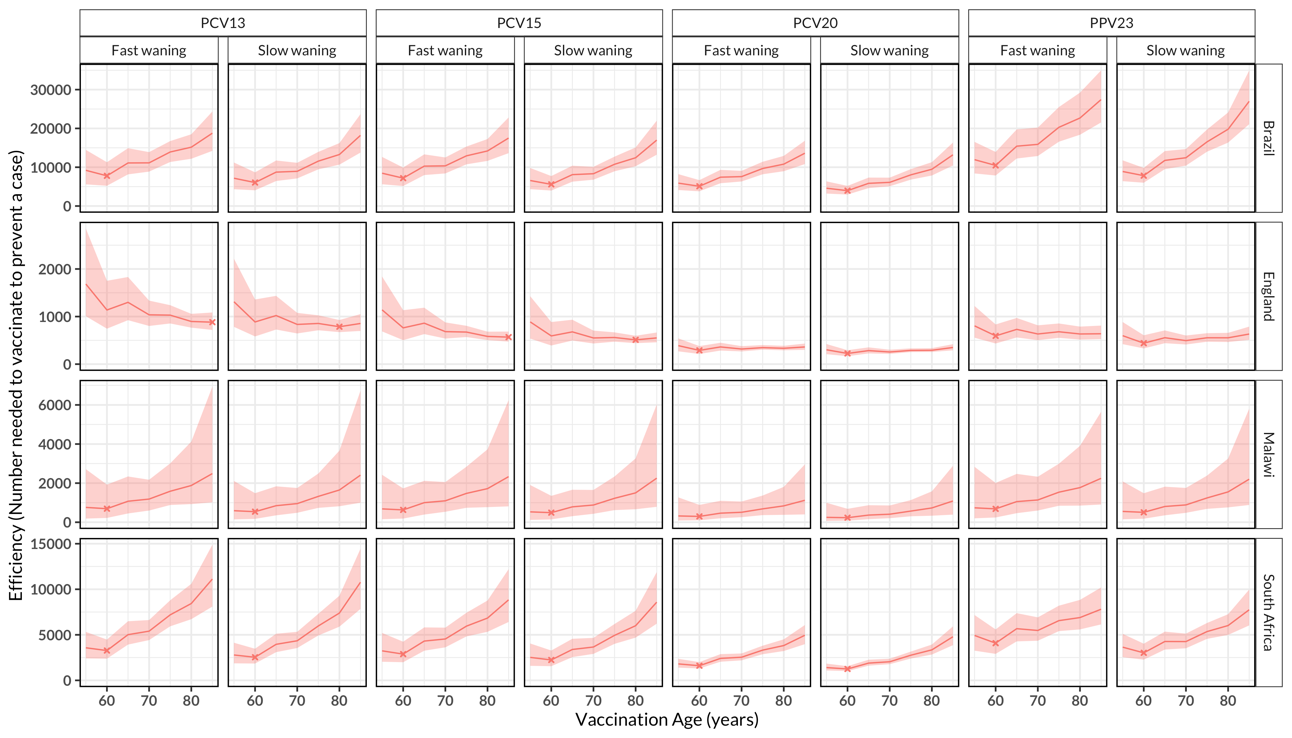

**Figure S7**. The efficiency of routine pneumococcal vaccination in older adults aged ≥55 years old (y). The number of individuals who are needed to vaccinate in each age of vaccination to prevent a case, stratified by country and vaccine product, under alternative scenario of age-dependent initial vaccine efficacy/effectiveness (VE) and waning VE in Brazil, England, Malawi and South Africa. The lines represent cohort model mean estimates and the shaded ribbon represents 95% bootstrap confidence intervals for the mean estimates. The X represents the optimal age for efficiency of pneumococcal vaccination program. In Brazil, Malawi and South Africa, optimal age of vaccination efficiency is achieved in 60y age cohort irrespective of vaccine product or waning VE assumption whereas in England, efficiency is achieved in 60y age cohort for PCV20 and PPV23 irrespective of waning VE assumption, and in 80y or 85y for PCV13 and PCV15’s slow or fast waning VE, respectively.

**References**

[1]	Brandileone M-CC, Almeida SCG, Minamisava R, Andrade A-L. Distribution of invasive Streptococcus pneumoniae serotypes before and 5 years after the introduction of 10-valent pneumococcal conjugate vaccine in Brazil. Vaccine 2018;36:2559–66. https://doi.org/10.1016/j.vaccine.2018.04.010.

[2]	Djennad A, Ramsay ME, Pebody R, Fry NK, Sheppard C, Ladhani SN, et al. Effectiveness of 23-Valent Polysaccharide Pneumococcal Vaccine and Changes in Invasive Pneumococcal Disease Incidence from 2000 to 2017 in Those Aged 65 and Over in England and Wales. EClinicalMedicine 2019;6:42–50. https://doi.org/10.1016/j.eclinm.2018.12.007.

[3]	Bar-Zeev N, Swarthout TD, Everett DB, Alaerts M, Msefula J, Brown C, et al. Impact and effectiveness of 13-valent pneumococcal conjugate vaccine on population incidence of vaccine and non-vaccine serotype invasive pneumococcal disease in Blantyre, Malawi, 2006–18: prospective observational time-series and case-control studies. Lancet Glob Health 2021;9:e989–98. https://doi.org/10.1016/S2214-109X(21)00165-0.

[4]	SanJoaquin MA, Allain TJ, Molyneux ME, Benjamin L, Everett DB, Gadabu O, et al. Surveillance Programme of IN-patients and Epidemiology (SPINE): Implementation of an Electronic Data Collection Tool within a Large Hospital in Malawi. PLOS Med 2013;10:e1001400. https://doi.org/10.1371/journal.pmed.1001400.

[5]	Meiring S, Cohen C, Quan V, Gouveia L de, Feldman C, Karstaedt A, et al. HIV Infection and the Epidemiology of Invasive Pneumococcal Disease (IPD) in South African Adults and Older Children Prior to the Introduction of a Pneumococcal Conjugate Vaccine (PCV). PLOS ONE 2016;11:e0149104. https://doi.org/10.1371/journal.pone.0149104.

[6]	Kleynhans J, Cohen C, McMorrow M, Tempia S, Crowther-Gibson P, Quan V, et al. Can pneumococcal meningitis surveillance be used to assess the impact of pneumococcal conjugate vaccine on total invasive pneumococcal disease? A case-study from South Africa, 2005–2016. Vaccine 2019;37:5724–30. https://doi.org/10.1016/j.vaccine.2019.04.090.

[7]	Srinivasan V, Mason CH. Nonlinear Least Squares Estimation of New Product Diffusion Models. Mark Sci 1986;5:169–78.
